# From Tabulated Data to Knowledge Graph: A Novel Way of Improving the Performance of the Classification Models in the Healthcare Data

**DOI:** 10.1101/2021.06.09.21258123

**Authors:** Nazar Zaki, Elfadil A Mohamed, Tetiana Habuza

## Abstract

In sectors like healthcare, having classification models that are both reliable and accurate is vital. Regrettably, contemporary classification techniques employing machine learning disregard the correlations between instances within data. This research, to rectify this, introduces a basic but effective technique for converting tabulated data into data graphs, incorporating structural correlations. Graphs have a unique capacity to capture structural correlations between data, allowing us to gain a deeper insight in comparison to carrying out isolated data analysis. The suggested technique underwent testing once the integration of graph data structure-related elements had been carried out and returned superior results to testing solely employing original features. The suggested technique achieved validity by returning significantly improved levels of accuracy.

**Data:** The extracted graph topological features datasets are available from:

## 1. Introduction

Over the last few years, there has been an explosion in the interest in the deployment of artificial intelligence, especially machine learning (ML) technologies for improving and accelerating the decision-making process within healthcare. This technology is seen as having the potential to provide diagnostic insights swiftly and accurately. A number of machine learning and deep learning (DL) methodologies have been created. These techniques have led to improvements in the accuracy of assessing diseases and fewer errors in treating disease. Nevertheless, numerous challenges still exist in terms of the development of workable ML models for use in healthcare. The difficulty of these challenges is exacerbated by the complex nature and sheer size of real-world data, particularly in the current Big Data environment. Additionally, numerous ML solutions, especially ones employing classification models, did not fulfill the hopes of the creators. The development of extremely accurate and workable classification models in a sector like healthcare is highly challenging. If it is shown to be possible to overcome problems concerning the quality of data, its volume, and complexity, more research is urgently needed regarding the way knowledge can be acquired and utilized practically within the healthcare sector [1].

In sectors like healthcare, this is vital that the classification model should be both reliable and accurate. Although researchers have recently used numerous high-quality and accurate classification methodologies, e.g., deep learning, not every classification model has demonstrated adequate superiority over previous techniques. This is due to the fact that these techniques gave little attention to correlations between data instances. In this research, it is demonstrated how the incorporation of basic knowledge graph algorithms can lead to improvements in the way classification models perform. Knowledge graph algorithms are created with a focus on correlations, and they have a unique capacity to discover structures and provide insights from connected data.

### 1.1. Knowledge Graphs in Healthcare

As ML and knowledge discovery have rapidly developed in recent times, numerous new forms of analysis of graphs and algorithm mining have been used in a number of areas. Healthcare is one such area, a sector that has been under significant pressure as a result of the COVID-19 pandemic. The development of ML models that can understand how diseases are transmitted, treated, and prevented is urgently needed; these models should be able to mine data from numerous sources, including academic and professional literature, hospital records, the pharmaceutical industry, and biological, microbiological, and genetic research. In order to do this, ML techniques incorporating analytics based on graphs are showing significant potential. It is not surprising that knowledge graphs have been widely used in the healthcare sector in recent times and we may categories the different types that can be applied as generative graph models, knowledge graph construction and inference, and network embedding.

### 1.2. Knowledge Graph Construction/Inference

Pham et al [2] have suggested the construction of knowledge-based heterogeneous information graphs to be used for classifications of medical health status. He et al [3] created synthetic triples using conceptualization, formulating the challenge as a triple classification that was addressed employing a discriminatory model, transferring knowledge from previously prepared language models. These researchers showed that their suggested methodology was effective in the identification of plausible triples and expansion of the knowledge graph using triples that could be highly diverse and novel in terms of edges and nodes. Additionally, Zhu et al [4] undertook a review of the literature regarding extant pharmaceutical knowledge bases and how they are applied in research by medical informatics.

### 1.3. Network Embedding

Yue et al. [5] undertook systematic comparisons with three significant predictive tasks for biomedical purposes: predictions of drug-disease Association (DDA), drug-drug interaction (DDI), and protein-protein interaction (PPI). They have provided a guideline framework for the proper selection of graph embedding methodology and delineating hyper-parameters for a variety of biomedical endeavors. Tu et al [6] put forward a Hyper-Network embedding model for the embedding of hyper networks having indecomposable hyperedges. This model was used with a quartet of different forms of hyper-network, including a drug network, and its performance showed promise. Chang Su et al. [7] undertook a review of the literature related to the application of networking embedding to effect advances within biomedicine. Baytas et al. [8] suggested a deep strategy for embedding heterogeneous attributed hyper-networks using complex non-linear node correlations. This involves the design of fully connected graph convolutional layers for the projection of a variety of node types.

### 1.4. Generative Graph Model Applications

Chengxi Zang et al. [9] created MoFlow, and an invertible flow model for the generation of molecular graphs. Ling Chen et al. [10] proposed a partially supervised learning algorithm based on graphs in order to mine data from health examinations and to predict risks for the classification of ongoing situations in which most of the data was not labeled. Sacchet MD et al. [11] employed a support vector machines classifier for distinguishing between individuals suffering from depression and those who were not on the basis of a number of brain network indications. These researchers also carried out an assessment of the value of specific graph metrics for the differentiation of the different classes.

Although these methods proved successful, a challenge is presented when faced with the unavailability of network data or if data is available but in standard tabulated format. To tackle this problem Tao et al. [1] designed a knowledge graph employing healthcare categorization and other knowledge mined from the NHANES data set, a collection of latent concepts employing the Pearson Correlation for decoding. This research presents a non-complex but extremely accurate way of converting tabulated data into graphs, allowing significant improvements in ML classifications.

## 2. Methodology

### 2.1. Data

The suggested technique underwent testing using well-regarded data sets in the healthcare field:

- D1 - Pima Indians Diabetes Dataset [12]: this is a dataset built by the National Institute of Diabetes and Digestive and Kidney Diseases. The database aims to provide diagnostic predictions as to the likelihood of a patient having diabetes on the basis of various diagnostic data including age in years, diabetes pedigree function, BMI (kilograms), insulin (mu U/m), skin thickness (millimeters) blood pressure (mm Hg), plasma glucose concentration levels, and the number of times pregnant. The dataset is publically available from Kaggle: https://www.kaggle.com/uciml/pima-indians-diabetes-database
- D2 - Stroke Prediction Dataset: this is a dataset containing a dozen attributes, these being patient id, gender, age in years, hypertension, heart_disease, ever_married, work_type, residence_type, avg_glucose_level, body mass index in kg), and smoking_status. There are missing entries in the data and setting attributes are normal, and so a pre-processing stage was required for imputation of missing entries and conversion of nominal attributes to integers. The dataset is publically available from Kaggle: https://www.kaggle.com/fedesoriano/stroke-prediction-dataset
- D3 - Heart Attack Analysis and Prediction Dataset [13]: this database is used for classifying heart attacks and has 14 data points including the likelihood of heart attack, maximum heart rate, resting ECG, fasting blood sugar (mg/dl), cholesterol level (mg/dl), blood pressure (mm Hg), type of chest pain, number of major vessels, angina induced by exercise, sex, and age (years). The dataset is publically available from Kaggle: https://www.kaggle.com/rashikrahmanpritom/heart-attack-analysis-prediction-dataset/metadata
- D4 - Hepatocellular Carcinoma dataset [14], [15]: this dataset was provided by the ML Repository, containing laboratory values from blood donors as well as patients with hepatocellular carcinoma, along with demographic information including gender and age. The dataset was obtained from UCI Machine Learning Repository: https://archive.ics.uci.edu/ml/datasets/HCV+data

Table 1 offers greater detail about the datasets employed by this research.

**Table 1:**
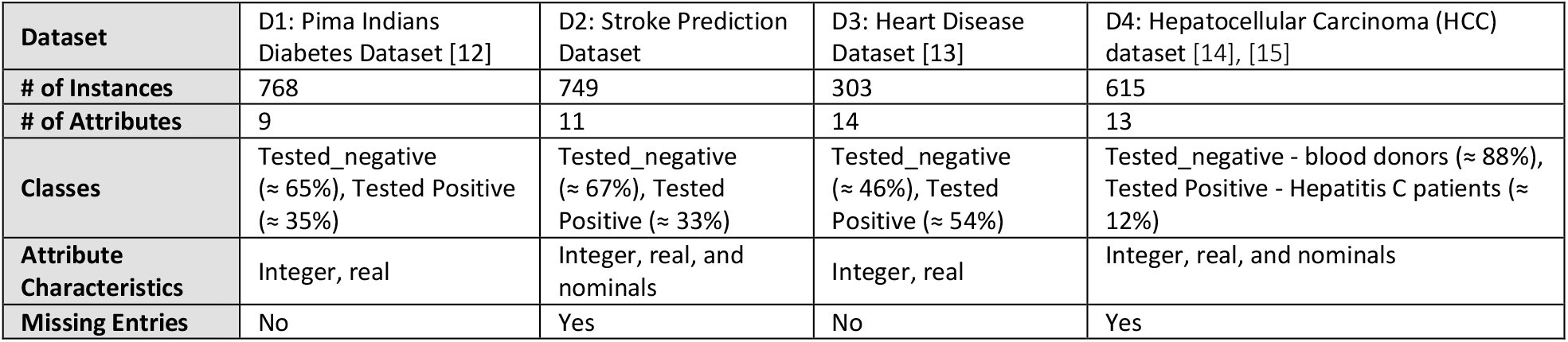
Datasets employed in this research

| Dataset | D1: Pima Indians Diabetes Dataset [12] | D2: Stroke Prediction Dataset | D3: Heart Disease Dataset [13] | D4: Hepatocellular Carcinoma (HCC) dataset [14], [15] |
| --- | --- | --- | --- | --- |
| # of Instances | 768 | 749 | 303 | 615 |
| # of Attributes | 9 | 11 | 14 | 13 |
| Classes | Tested_negative ($\approx 65\%$), Tested Positive ($\approx 35\%$) | Tested_negative ($\approx 67\%$), Tested Positive ($\approx 33\%$) | Tested_negative ($\approx 46\%$), Tested Positive ($\approx 54\%$) | Tested_negative - blood donors ($\approx 88\%$), Tested Positive - Hepatitis C patients ($\approx 12\%$) |
| Attribute Characteristics | Integer, real | Integer, real, and nominals | Integer, real | Integer, real, and nominals |
| Missing Entries | No | Yes | No | Yes |

Figure 1 offers an overview of the suggested methodology.

**Figure 1:**
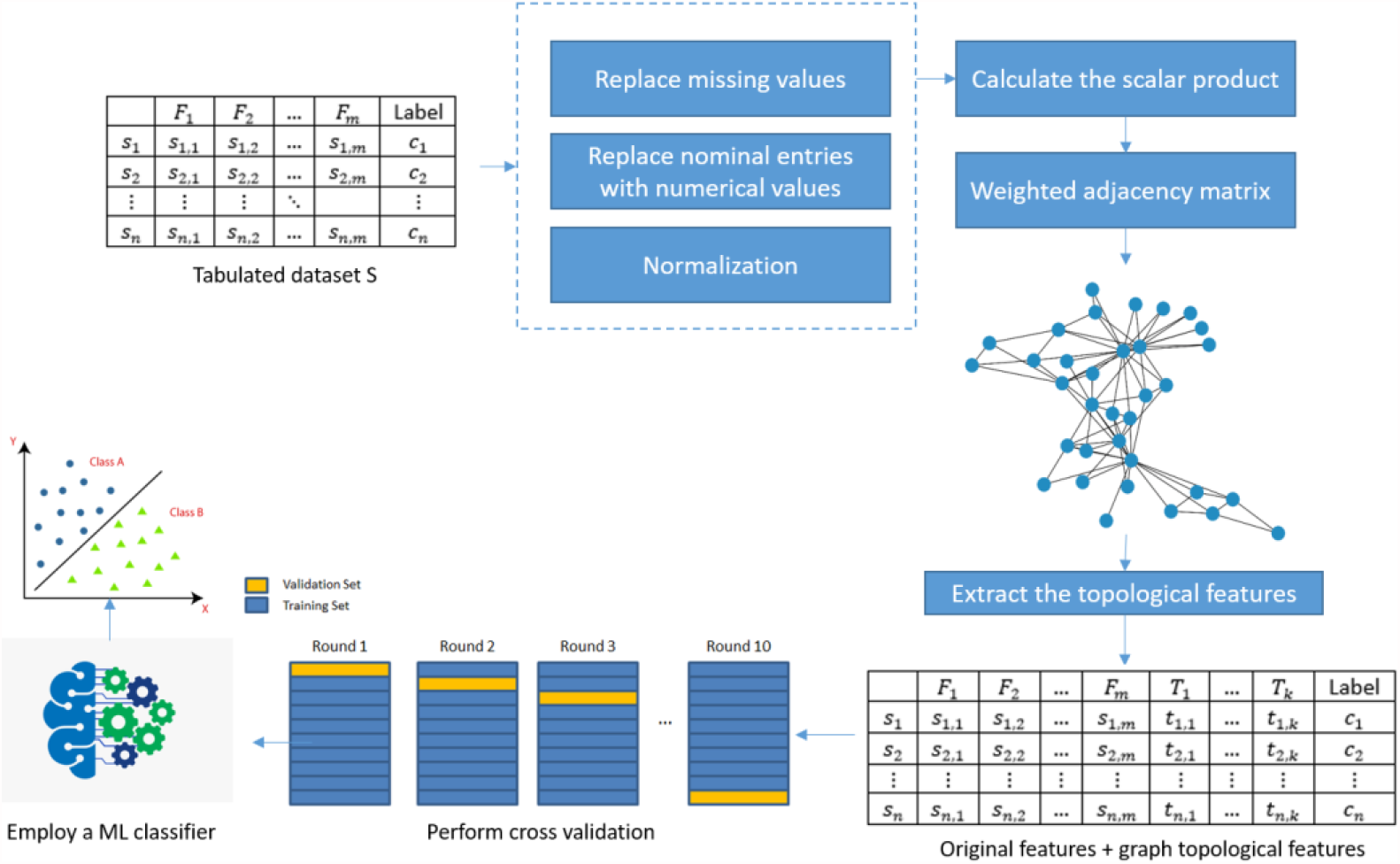
Overview of the suggested methodology

For illustration of the methodology, an assumption is made that there is a hypothetical dataset *S* of *n* samples 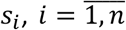. Every sample *s*_*i*_ is shown through *m* features 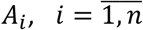. For simplifying the difficulty, an assumption is made that every feature is defined, i.e. *A*_*i*_ ∈ ℝ. A typical dataset structure would be:

| | $F_1$ | $F_2$ | ... | $F_m$ | Label |
| --- | --- | --- | --- | --- | --- |
| $s_1$ | $s_{1,1}$ | $s_{1,2}$ | ... | $s_{1,m}$ | $c_1$ |
| $s_2$ | $s_{2,1}$ | $s_{2,2}$ | ... | $s_{2,m}$ | $c_2$ |
| ⋮ | ⋮ | ⋮ | $\ddots$ | | ⋮ |
| $s_n$ | $s_{n,1}$ | $s_{n,2}$ | ... | $s_{n,m}$ | $c_n$ |

Every sample *s*_*i*_ in the dataset, *S* is part of to class *c*_*i*_, where *c*_*i*_ ∈ *C, C* = {*C*_1_, *C*_2_, … *C*_*l*_}. This is a standard *l*-class classification problem that may be resolved employing ML methods. For integrating graph-related features, we must firstly use the scalar product operation, thus:

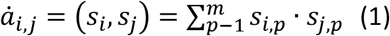

In this instance the adjacency matrix 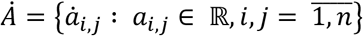, is representative of a weighted graph *G* having *n* nodes. Additionally, we employ a threshold technique for further simplification of the problem, e.g., if 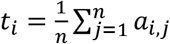, the adjacency matrix A of unweighted G (see below) can be written as

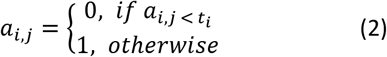

| | $s_1$ | $s_1$ | ... | $s_n$ | Label |
| --- | --- | --- | --- | --- | --- |
| $s_1$ | 0 | 0\|1 | ... | 0\|1 | $c_1$ |
| $s_1$ | 0\|1 | 0 | ... | 0\|1 | $c_2$ |
| ⋮ | ⋮ | ⋮ | $\ddots$ | | ⋮ |
| $s_n$ | 0\|1 | 0\|1 | ... | 0 | $c_n$ |

For the extraction of meaningful features, we can represent graph G’s notes employing the topological features 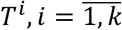, a few of which are PageRank value, centrality score, and degree of the node. These topological features can then be integrated with original features for incorporation of useful graph information and finding correlations between instances, thus:

| | $F_1$ | $F_2$ | ... | $F_m$ | $T_1$ | ... | $T_k$ | Label |
| --- | --- | --- | --- | --- | --- | --- | --- | --- |
| $s_1$ | $s_{1,1}$ | $s_{1,2}$ | ... | $s_{1,m}$ | $t_{1,1}$ | ... | $t_{1,k}$ | $c_1$ |
| $s_2$ | $s_{2,1}$ | $s_{2,2}$ | ... | $s_{2,m}$ | $t_{2,1}$ | ... | $t_{2,k}$ | $c_2$ |
| ⋮ | ⋮ | ⋮ | ⋮ | ⋮ | ⋮ | ⋮ | ⋮ | ⋮ |
| $s_n$ | $s_{n,1}$ | $s_{n,2}$ | ... | $s_{n,m}$ | $t_{n,1}$ | ... | $t_{n,k}$ | $c_n$ |

In this instance, the matrix T may be made up of the previously noted features, thus:

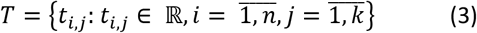

The topological features under consideration in this research can be seen in summary in Table 2: Topological features extracted from the pre-processed datasetTable 2. The features final extracted can then be employed alongside the ML classifier, which will provide classifications of greater accuracy.

**Table 2:** Topological features extracted from the pre-processed dataset

| # | Feature | Definition | Formula |
| --- | --- | --- | --- |
| 1 | Degrees | The number of edges connected to a node | $deg(s_i) = 2 \cdot \sum_{j=1}^n a_{i,j}$ |
| 2 | In degree | The number of head ends adjacent to a node | $deg^-(s_i) = \sum_{j=1}^n a_{j,i}$ |
| 3 | Out degree | The number of tail ends adjacent to a node | $deg^+(s_i) = \sum_{j=1}^n a_{i,j}$ |
| 4 | Eccentricity | Maximum distance between a node and any other node in the graph | $e(s_i) = \max(d(s_i, s_1), d(s_i, s_2), \dots, d(s_i, s_n))$ |
| 5 | Weighted degree | The summation of edges connected to a node | $deg^{weighted}(s_i) = deg^-(s_i) + deg^+(s_i)$ |
| 6 | Hub | The number of highly authoritative nodes this node is pointing to | $h(s_i) = \sum_{s_i \mapsto y} a(y)$. Initially, $h(s_i) = 1$ |
| 7 | Authority | The amount of valuable information that a node holds | $a(s_i) = \sum_{y \mapsto s_i} a(y)$. Initially, $a(s_i) = 1$ |
| 8 | PageRank | A measure of the importance of a node within the graph | $PR(s_i) = \frac{1-c}{n} + c \cdot \sum_{s_j \in M(s_i)} \frac{PR(s_j)}{deg^+(s_j)}$, $c = 0.85$ |
| 9 | Triangle | A number of triangles that include a node $s_i$ as one vertex | $tr(s_i) = \sum_{s_j, s_k \in M(s_i)} \{s_j, s_k\}$ |
| 10 | Closeness centrality | Time it takes to reach other nodes in the graph | $C(s_i) = \frac{1}{\sum_{j=1}^n d(s_i, s_j)}$ |
| 11 | Betweenness Centrality | An indicator of a node centrality or importance in the graph | $C_B(s_i) = \sum_{s_i \neq s_j \neq s_p} \frac{\sigma_{s_i, s_p}(s_j)}{\sigma_{s_j, s_p}}$ |
| 12 | Harmonic Centrality | Allows to reverse the sum and reciprocal operations in closeness centrality | $C_H(s_i) = \sum_{j=1}^n \frac{1}{d(s_i, s_j)}, j \neq i$ |
| 13 | Eigenvector centrality | A measure of the transitive influence or connectivity of nodes | $x(s_i) = \frac{1}{\lambda} \sum_{j=1}^n a_{i,j} \cdot x(s_j)$ |

\* $\{s_i, s_j\}$ is an edge between nodes $s_i$ and $s_j$ .
$d(s_i, s_j)$ is a distance between nodes $s_i$ and $s_j$ .
$s_i \mapsto y$ denotes an existence of a link between $s_i$ and $y$ nodes.
$\sigma_{s_j, s_p}$ is a total number of shortest paths from node $s_j$ to node $s_p$ , $j, p = \overline{1, n}$ . $\sigma_{s_i, s_p}(s_i)$ the number of those paths that pass through $s_i$ .
$M(s_i)$ is a set of nodes that has link to $s_i$ .

### 2.2. Evaluation Measures

For evaluating the performance of the suggested methodology, we employed 10-fold cross-validation. Widely excepted evaluation measures were employed for evaluation of the accuracy of the classifications, including F1-score, Recall (RE), Precision (PR), and Accuracy (AC).

## 3. Experimental work/results

This section will detail the experimental work undertaken for effecting significant improvements in the accuracy of classifications through the conversion of tabulated data structures into graph data structures. It is predicted that graphs will be able to capture extra significant correlations between instances that are frequently disregarded in the process of classification. The intention is to undertake an evaluation of the benefits of the incorporation of adding graph-related features to original features. It is a simple matter to calculate the accuracy of classifications either with or without the suggested graph additions.

In order to test this new methodology, we began by preprocessing the data sets, ensuring that numerical values replaced nominal entries and that every missing entry was imputed (employing simple methodologies, e.g. average values). Normalization of the dataset values was then undertaken using a standard scaler or the MinMax scaler. In the initial phase, we employed a number of classifiers, including Naïve Bayes (NB), neural network (NN), support vector machines (SVM), logistic regression (LR), k-nearest neighbor (KNN), random forest (RF), and decision tree (DT) forecast for classifying both classes in all four data sets and recording the accuracy of classification. We only considered the original features (OF) and default parameter values. In the second phase, we only considered topological features. Additionally, we also tested combinations of original features and data extracted from the graphs. As Table 3 shows, incorporating the graph topological features affected a significant improvement in classification accuracy for every dataset and every classifier.

**Table 3:** Summary of classification model performance on the basis of original features, graph topological features, and a combination of the two with each dataset. Seven classification techniques are employed for measuring classification performance.

|  |  | D1: Pima Indians Diabetes Dataset |  |  | D2: Stroke Prediction Dataset |  |  | D3: Heart Disease Dataset |  |  | D4: Hepatocellular Carcinoma (HCC) dataset |  |  |
| --- | --- | --- | --- | --- | --- | --- | --- | --- | --- | --- | --- | --- | --- |
|  |  | OF* | GTF* | OF+GTF | OF* | GTF* | OF+GTF | OF* | GTF* | OF+GTF | OF* | GTF* | OF+GTF |
| NB | Accuracy | 0.76 | 0.85 | 0.76 | 0.76 | 0.9 | 0.9 | 0.83 | 0.93 | 0.96 | 0.93 | 0.92 | 0.95 |
| NB | Precision | 0.76 | 0.85 | 0.84 | 0.77 | 0.90 | 0.90 | 0.83 | 0.93 | 0.96 | 0.93 | 0.93 | 0.95 |
| NB | Recall | 0.76 | 0.85 | 0.84 | 0.76 | 0.90 | 0.90 | 0.83 | 0.93 | 0.96 | 0.93 | 0.92 | 0.95 |
| NB | F1 score | 0.76 | 0.85 | 0.84 | 0.76 | 0.90 | 0.90 | 0.83 | 0.93 | 0.96 | 0.93 | 0.92 | 0.95 |
| NN | Accuracy | 0.75 | 0.98 | 0.98 | 0.72 | 0.98 | 0.97 | 0.78 | 0.97 | 0.98 | 0.96 | 0.97 | 0.98 |
| NN | Precision | 0.75 | 0.98 | 0.98 | 0.72 | 0.98 | 0.97 | 0.78 | 0.97 | 0.98 | 0.96 | 0.97 | 0.98 |
| NN | Recall | 0.75 | 0.98 | 0.98 | 0.72 | 0.98 | 0.97 | 0.78 | 0.97 | 0.98 | 0.96 | 0.97 | 0.98 |
| NN | F1 score | 0.75 | 0.98 | 0.98 | 0.72 | 0.98 | 0.97 | 0.78 | 0.97 | 0.98 | 0.96 | 0.97 | 0.98 |
| SVM | Accuracy | 0.77 | 0.95 | 0.95 | 0.76 | 0.96 | 0.97 | 0.84 | 0.97 | 0.98 | 0.93 | 0.96 | 0.97 |
| SVM | Precision | 0.77 | 0.95 | 0.96 | 0.77 | 0.96 | 0.97 | 0.84 | 0.97 | 0.98 | 0.92 | 0.96 | 0.97 |
| SVM | Recall | 0.77 | 0.95 | 0.95 | 0.76 | 0.96 | 0.97 | 0.84 | 0.97 | 0.98 | 0.93 | 0.96 | 0.97 |
| SVM | F1 score | 0.77 | 0.95 | 0.95 | 0.76 | 0.96 | 0.97 | 0.84 | 0.97 | 0.98 | 0.92 | 0.96 | 0.97 |
| LR | Accuracy | 0.77 | 0.98 | 0.97 | 0.77 | 0.98 | 0.98 | 0.82 | 0.97 | 0.96 | 0.96 | 0.99 | 0.98 |
| LR | Precision | 0.76 | 0.98 | 0.97 | 0.77 | 0.98 | 0.98 | 0.82 | 0.97 | 0.96 | 0.96 | 0.99 | 0.98 |
| LR | Recall | 0.77 | 0.98 | 0.97 | 0.77 | 0.98 | 0.98 | 0.82 | 0.97 | 0.96 | 0.96 | 0.99 | 0.98 |
| LR | F1 score | 0.77 | 0.98 | 0.97 | 0.77 | 0.98 | 0.98 | 0.82 | 0.97 | 0.96 | 0.96 | 0.99 | 0.98 |
| KNN | Accuracy | 0.71 | 0.97 | 0.93 | 0.68 | 0.96 | 0.93 | 0.77 | 0.98 | 0.93 | 0.92 | 0.97 | 0.96 |
| KNN | Precision | 0.71 | 0.97 | 0.93 | 0.68 | 0.96 | 0.93 | 0.77 | 0.98 | 0.93 | 0.91 | 0.97 | 0.96 |
| KNN | Recall | 0.72 | 0.97 | 0.93 | 0.68 | 0.96 | 0.93 | 0.77 | 0.98 | 0.93 | 0.92 | 0.97 | 0.96 |
| KNN | F1 score | 0.71 | 0.97 | 0.93 | 0.68 | 0.96 | 0.93 | 0.77 | 0.98 | 0.93 | 0.91 | 0.97 | 0.96 |
| RF | Accuracy | 0.76 | 0.97 | 0.97 | 0.74 | 0.97 | 0.97 | 0.82 | 0.98 | 0.99 | 0.98 | 0.98 | 0.98 |
| RF | Precision | 0.76 | 0.97 | 0.97 | 0.74 | 0.97 | 0.97 | 0.82 | 0.98 | 0.99 | 0.98 | 0.98 | 0.98 |
| RF | Recall | 0.76 | 0.97 | 0.97 | 0.74 | 0.97 | 0.97 | 0.82 | 0.98 | 0.99 | 0.98 | 0.98 | 0.98 |
| RF | F1 score | 0.76 | 0.97 | 0.97 | 0.74 | 0.97 | 0.97 | 0.82 | 0.98 | 0.99 | 0.98 | 0.98 | 0.98 |
| DT | Accuracy | 0.71 | 0.96 | 0.95 | 0.73 | 0.96 | 0.97 | 0.79 | 0.99 | 0.99 | 0.96 | 0.98 | 0.97 |
| DT | Precision | 0.71 | 0.96 | 0.96 | 0.73 | 0.96 | 0.97 | 0.79 | 0.99 | 0.99 | 0.96 | 0.98 | 0.97 |
| DT | Recall | 0.71 | 0.96 | 0.95 | 0.73 | 0.96 | 0.97 | 0.79 | 0.99 | 0.99 | 0.96 | 0.98 | 0.97 |
| DT | F1 score | 0.71 | 0.96 | 0.95 | 0.73 | 0.96 | 0.97 | 0.79 | 0.99 | 0.99 | 0.96 | 0.98 | 0.97 |

\*OF – Original features
\*GTF – Graph topological features

Lastly, an elementary correlation-based feature subset selection [16] was undertaken for identification of an elementary correlation-based feature subset selection [16] was undertaken for identification of extremely informative features (either original or graph topological) and the measurement of the accuracy with a limited feature set. As table 4 shows, although 18% of the features based on D1 were selected, 21% of D2, 30% of D3, and 42% of D4, classification accuracy can be compared to the case when we consider every feature (original and graph topological).

**Table 4:**
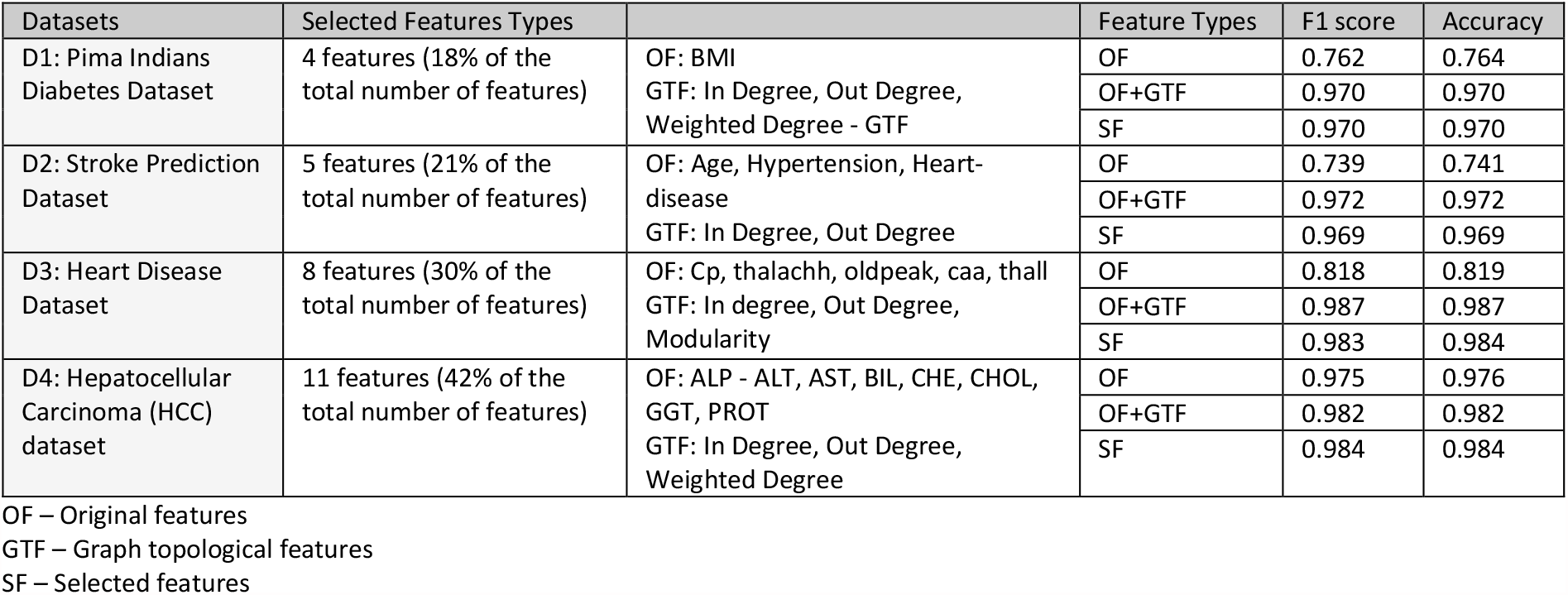
Classification performance when employing OF, combining OF and GTP, with the selected features

We additionally undertook an investigation of the information offered by every feature in order to increase the accuracy of classification. We employed heat map visualization techniques to illustrate the level that each feature adds to the classification model in 2D color. Figure 2: heat map visualization demonstrates that the graph’s topological features have a high level of correlation with class attributes, not original attributes; in each of the four datasets the topological features demonstrated more correlation with the class.

**Figure 2:**
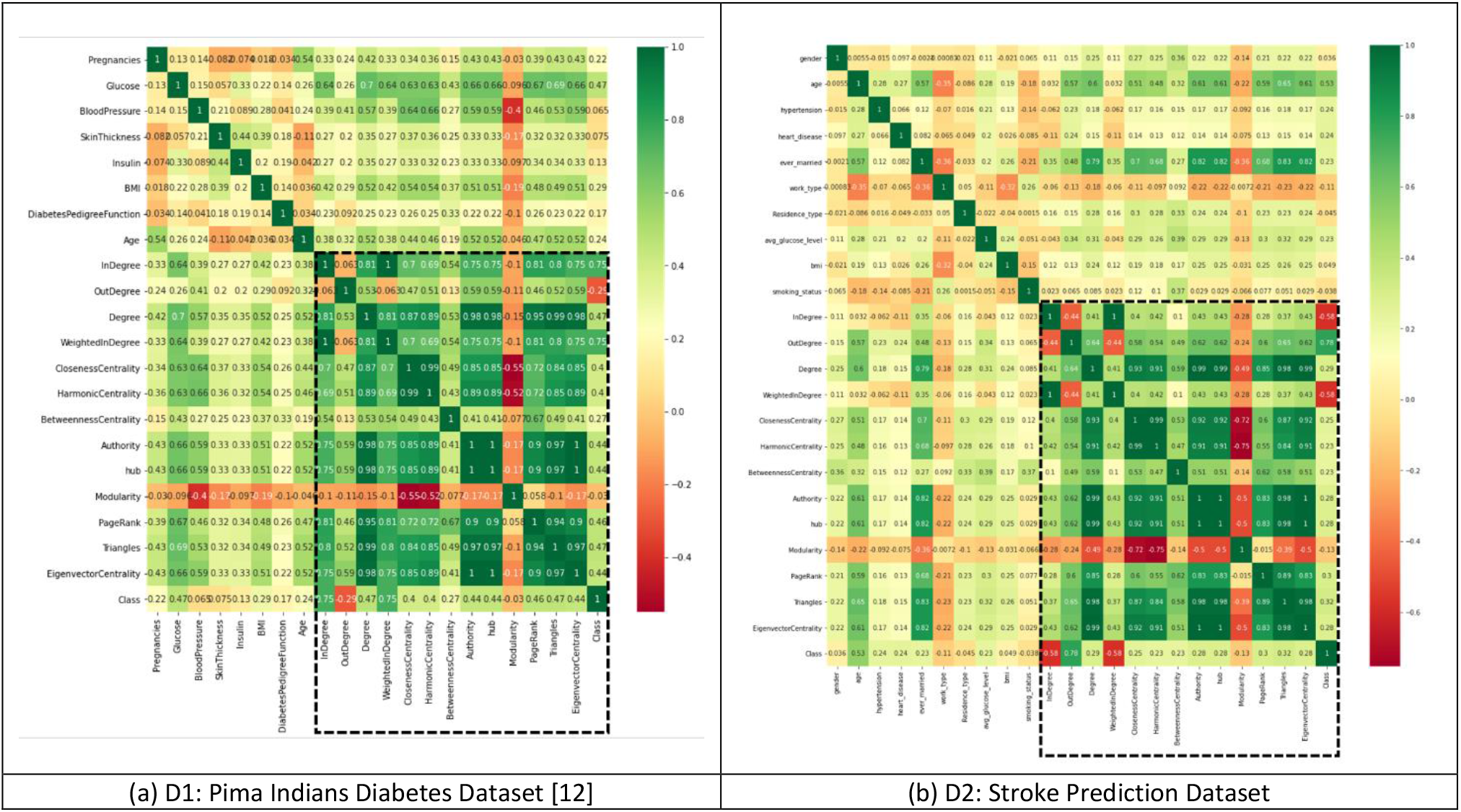
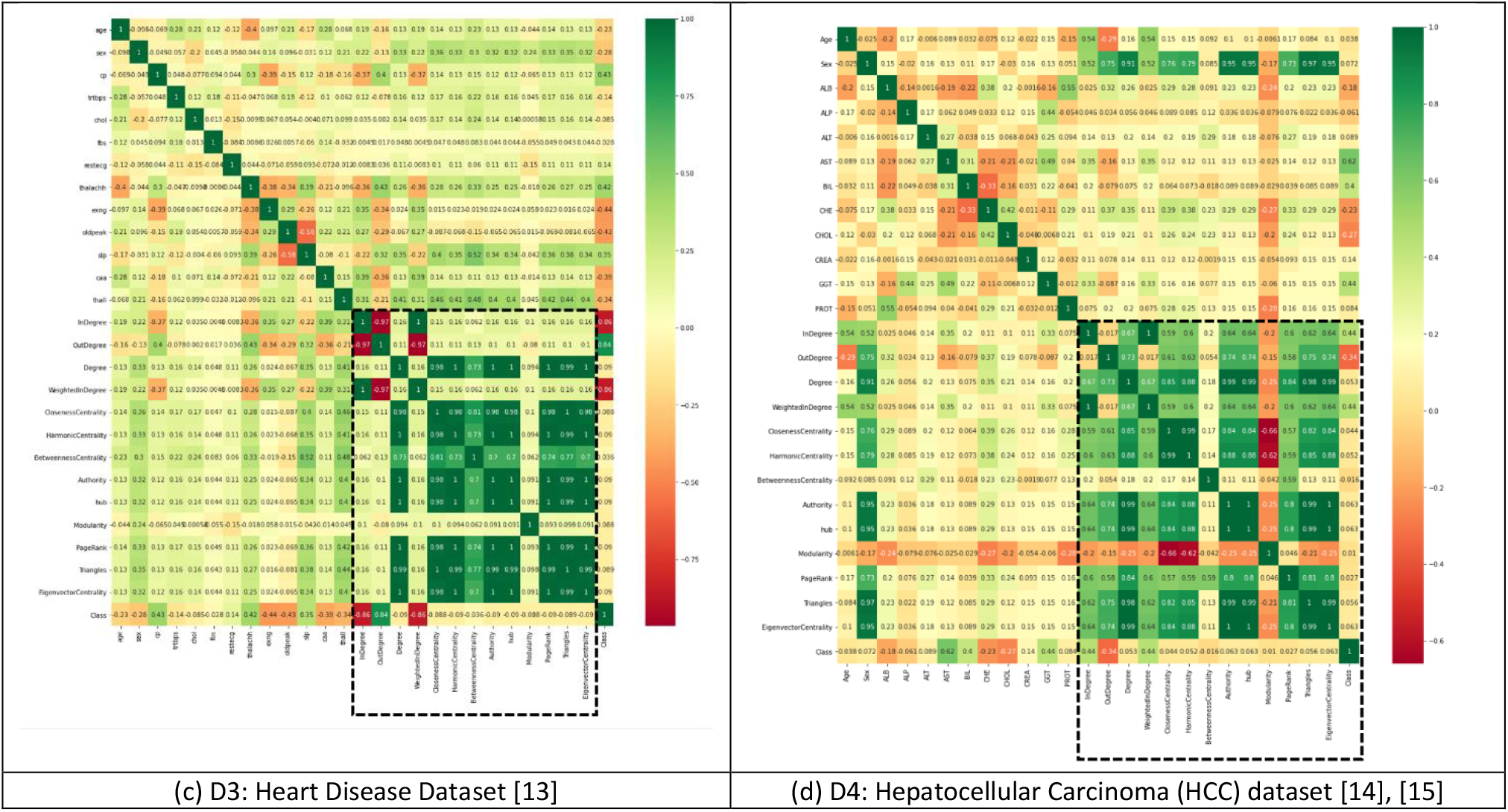
Heat map visualization demonstrating that graphs of topological features have a high degree of correlation with class attributes, not original attributes.

## 4. Conclusion/discussion

Graphs have a unique capacity for the capture of structural correlations between data and this provides greater insight in comparison to isolated data analysis [17]. Many forms of data are not formulated as graphs originally, e.g., text data, images, and tabulated data. In this research, we have put forward a basic methodology for the conversion of tabulated data to graph data. Whilst this methodology is relatively simple, it has been shown to be a powerful new way of interpreting data. As the result section demonstrates, there have been significant improvements to the classification performance with all of the four sample data sets. Incorporating graph topological features has added information to the classifier which has resulted in better performance. The classifications may be improved to an even greater extent by applying more powerful classification techniques, e.g. deep learning. We did not test deep-learning-based classifiers in this research as the data sets are relatively small. Additionally, using a convolutional graph network (GCN) with the features extracted may return promising results. GCN has recently come to the fore in ML and related disciplines and has shown that it can provide improved results in a number of areas. Although our proposed methodology was tested in a healthcare context, it is a relatively generic methodology that can be employed for improving the classification accuracy of tabulated data in all disciplines. One limitation of the suggested methodology is that it will only work with numerical data. This meant that in this research nominal data had to be transformed to a numerical value when working with the D2 and D4 databases.

## Supporting information

Topological features

## Data Availability

All data sets used in this study are publically available online either from Kaggle (https://www.kaggle.com/) or UCI Machine Learning Repository (https://archive.ics.uci.edu/ml/datasets/)

D1-Pima Indians Diabetes Dataset: https://www.kaggle.com/uciml/pima-indians-diabetes-database

D2-Stroke Prediction Dataset: https://www.kaggle.com/fedesoriano/stroke-prediction-dataset

D3-Heart Attack Analysis and Prediction Dataset: https://www.kaggle.com/rashikrahmanpritom/heart-attack-analysis-prediction-dataset/metadata

D4-Hepatocellular Carcinoma dataset: https://archive.ics.uci.edu/ml/datasets/HCV+data

https://www.kaggle.com/uciml/pima-indians-diabetes-database

https://www.kaggle.com/fedesoriano/stroke-prediction-dataset

https://www.kaggle.com/rashikrahmanpritom/heart-attack-analysis-prediction-dataset/metadata

https://archive.ics.uci.edu/ml/datasets/HCV+data

## Abbreviations

ML: Machine learning
DL: Deep learning
DDA: Drug-disease association
DDI: Drug-drug interaction
LR: Logistic regression
SVM: Support vector machines
RF: Random forest
NN: Neural network
NB: Naïve Bayes
KNN: k-nearest neighbor
DT: Decision tree
GCN: Convolutional graph network
OF: Original features
GTF: Graph topological features
SF: Selected features

## Acknowledgment

The authors would like to acknowledge partial support from the big data analytics center, united Arab Emirates University. Special thanks to Mrs. Asha Plackal for her valuable help.

